# Impact of Medicaid Expansion on Lung Cancer Survival Outcomes: A Difference-in-Differences Analysis

**DOI:** 10.1101/2025.08.31.25334804

**Authors:** Oluwasegun Akinyemi, Mojisola Fasokun, Akachukwu Eze, Nkemdirim Ugochukwu, Sumaiyya Arshad, Orimisan Belie, Kakra Hughes, Edward Cornwell, Gal Levy

## Abstract

**INTRODUCTION:** The Affordable Care Act’s Medicaid expansion aimed to enhance healthcare access for low-income individuals and minority groups, promoting early screening and treatment to improve health equity.

**OBJECTIVE:** This study examines the impact of Medicaid expansion on lung cancer-specific survival (CSM) and overall mortality (OS) by comparing outcomes in Texas (non-expansion of ACA) and California (expansion of ACA).

**METHODOLOGY:** We conducted a retrospective study using data from SEER cancer registry (2007–2021) to evaluate the impact of Medicaid expansion on lung cancer survival in California (expansion) vs. Texas (non-expansion). The study included adults aged 18–64, with periods split into pre-ACA (2007–2013), one-year washout (2014), and post-ACA (2015–2021). We utilized a DID design and adjusted for important covariates.

**RESULTS:** Among 119,937 individuals with Lung cancer, 52.1% were in California (62,521), while 47.8% were in Texas (57,416). The pre-ACA period included 60,010 individuals (53.1% in California and 46.9% in Texas), and 59,927 patients were in the post-ACA period (51.2% in California and 48.8% in Texas). Overall, Medicaid expansion was associated with a 1.12-point (− 1.12, 95% CI –1.46 to –0.77) reduction in the hazard of cancer-specific mortality. The policy was also associated with a 0.81point reduction in the hazard of overall mortality (−0.81, 95% CI –1.06 to –0.57).

**CONCLUSION:** Medicaid expansion was associated with a significant improvement in lung cancer outcomes among individuals with lung cancer in California, which implemented the policy in 2014, compared to Texas, which has not yet implemented the policy.

## Introduction

Lung cancer remains a significant public health challenge in the United States, with an estimated 238,340 new cases diagnosed and 127,070 deaths expected in 2023[1–3]. It is the second most common cancer among both men and women, yet it accounts for the highest number of cancer-related deaths, surpassing breast, prostate, and colorectal cancers combined[3, 4]. The burden of lung cancer mortality is particularly striking, with men exhibiting slightly higher death rates than women[5, 6]. This disease not only claims lives but also imposes a substantial financial toll on the U.S. healthcare system, with direct medical costs exceeding $13 billion annually, which includes the initial phase and continuing care phase[7]. Beyond the healthcare expenses, lung cancer results in significant indirect costs, including billions of dollars in lost productivity due to premature deaths and work absences, highlighting the extensive economic impact of this disease on society[8–10].

Social determinants of health (SDOH) play a critical role in influencing lung cancer outcomes[11–13]. Disparities in mortality rates are pronounced across racial, ethnic, and socioeconomic lines, with the highest burden observed among Black, Hispanic, and low-income populations[14, 15]. Socioeconomically disadvantaged individuals face barriers to early detection and access to timely, effective treatment, exacerbating poor outcomes[15]. These disparities underscore the intersection of healthcare access, income inequality, and systemic racism in shaping health inequities, particularly for lung cancer, a disease with significant morbidity and mortality.

The Affordable Care Act (ACA), signed into law in 2010, sought to address these disparities through sweeping healthcare reforms aimed at increasing insurance coverage, reducing healthcare costs, and improving population health[16, 17]. One of the ACA’s major components is Medicaid expansion, a policy designed to extend health coverage to low-income adults[18]. The staggered implementation of Medicaid expansion across states provides a natural experiment to assess its impact on health outcomes. By reducing financial barriers and increasing access to preventive services and timely treatment, Medicaid expansion has the potential to improve outcomes for cancers such as lung cancer, which require early and continuous care for optimal management.

In California, Medicaid expansion, implemented in 2014 under the Medi-Cal program, aimed to increase healthcare access among low-income residents. The state’s decision to expand Medicaid was part of a broader strategy to address healthcare inequities and improve outcomes for its diverse population[19]. Studies from states that adopted Medicaid expansion have reported reductions in cancer-related mortality, increased early-stage diagnoses, and improved access to treatment[20, 21]. These findings suggest that Medicaid expansion may play a crucial role in mitigating the burden of lung cancer, particularly among underserved populations.

This study aims to evaluate the impact of Medicaid expansion on lung cancer outcomes in California compared to Texas, a non-expansion state. By examining differences in cancer-specific and overall mortality between these states, this study seeks to provide insights into the role of Medicaid expansion in reducing disparities and improving survival for individuals diagnosed with lung cancer.

## Methodology

### Study Design

This retrospective comparative cohort study utilized population-based cancer registry data from the SEER registry[22]. SEER provides comprehensive data on cancer incidence, treatment, and outcomes, covering approximately 48% of the U.S. population[22, 23].

### Study Population

The study population consisted of individuals aged 18 to 64 years diagnosed with lung cancer between January 1, 2007, and December 31, 2021. Two states were selected for comparison: California, representing a Medicaid expansion state, and Texas, representing a non-expansion state. This design facilitated an evaluation of the impact of Medicaid expansion on lung cancer outcomes, focusing on cancer-specific mortality and overall mortality.

### Study Period and Periods of Interest

The study analyzed two distinct time periods: pre-ACA (2007–2013) and post-ACA (2015– 2021). A washout period in 2014 allowed for the full implementation of Medicaid expansion in California, which adopted the policy in 2014 under Medi-Cal.

### California vs. Texas

California implemented Medicaid expansion as part of the ACA in 2014, significantly increasing healthcare access for low-income populations through Medi-Cal. This program aimed to reduce uninsured rates, improve early detection, and enhance access to treatments, particularly for underserved populations. In contrast, Texas did not adopt Medicaid expansion, maintaining higher uninsured rates and potentially more limited access to care. These contrasting implementation statuses allowed for a comparative analysis of outcomes under different policy environments.

### Primary Outcomes of Interest

The primary outcomes of interest were cancer-specific mortality (CSM) and overall mortality (OM). Additionally, the results were stratified by income level, race/ethnicity, and disease stage at presentation to assess disparities and differential effects of Medicaid expansion.

### Cancer-Specific Mortality

(CSM) served as a crucial indicator of treatment effectiveness and long-term prognosis among lung cancer patients. Cox proportional hazards regression was used to examine the association between cancer-specific deaths and predictor variables, including state (California vs. Texas), period (pre-ACA vs. post-ACA), and their interaction.

### Overall Mortality

(OM) was used as a measure of general prognosis and included deaths from any cause. Cox regression analysis assessed the relationship between OM and predictor variables, similar to the analysis for CSM. These models enabled the evaluation of how Medicaid expansion influenced overall survival.

### Disease Stage at Presentation

The disease stage at diagnosis was categorized into three groups using SEER’s Combined Summary Stage system: localized, regional, and distant. This variable allowed consistent stage analysis over time.

Localized: Cancer confined to the primary site.

Regional: Cancer spread to regional lymph nodes or adjacent tissues.

Distant: Cancer spreads to remote organs or lymph nodes.

### Independent Variable of Interest

The primary explanatory variable was Medicaid expansion under the ACA. This variable was operationalized as a categorical variable representing two time periods: pre-ACA (2007–2013) and post-ACA (2015–2021). The interaction between the time period and state was used to evaluate the differential impact of Medicaid expansion on lung cancer outcomes.

### Covariates

Covariates included demographic factors such as age at diagnosis (18-44, 45-64 years), race/ethnicity (non-Hispanic White, non-Hispanic Black, Hispanic, Asian/Pacific Islander), and income level (<$65,000 and ≥$65,000). Clinical variables included disease stage at presentation (localized, regional, distant) and treatment modalities (surgery, chemotherapy, radiation).

### Theoretical Model: Andersen Behavioral Model

This study applied the Andersen Behavioral Model of Health Services to explore the influence of predisposing, enabling, and need factors on lung cancer outcomes. Predisposing factors included demographics (age, race, marital status); enabling factors encompassed income, metropolitan status, and Medicaid expansion status (California vs. Texas); and need factors reflected disease severity and treatment receipt.

### DID Specification

A DID model was used to estimate the impact of Medicaid expansion on lung cancer outcomes. The variable ACA was set to 1 for the post-ACA period (2015–2021) and 0 for the pre-ACA period (2007–2013). The variable State was defined as 1 for California (expansion state) and 0 for Texas (non-expansion state). The interaction term (ACA x State) provided the key estimate of Medicaid expansion’s impact.

The DID model is specified as[24]:

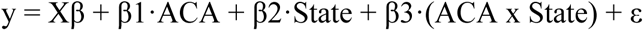

Where:

y represents the lung cancer outcome of interest (CSM or OM).
X includes covariates such as age, race, income, and treatment modalities.
β1 captures differences in outcomes across the pre- and post-ACA periods.
β2 captures baseline differences between California and Texas.
β3 represents the effect of Medicaid expansion.

To ensure the validity of our DiD analysis, we assessed the parallel trends assumption by comparing cancer-specific survival over time in Texas (non-expansion state) and California (expansion state) using Kaplan-Meier survival curves in both the pre-ACA (2007-2013) and post-ACA periods (2015-2021). In the pre-ACA period, survival trends were nearly identical between the two states, with no statistically significant difference, confirming that both groups followed similar survival trajectories before Medicaid expansion. However, in the post-ACA period, California exhibited a significant improvement in cancer-specific survival compared to Texas, supporting the hypothesis that Medicaid expansion contributed to enhanced survival outcomes (See Appendix: Figures 1& 2). These findings indicate that the parallel trends assumption was met, ensuring that the observed post-ACA survival differences can be attributed to Medicaid expansion rather than pre-existing differences between states. The corresponding Kaplan-Meier survival curves are presented as a supplementary file.

### Statistical Analysis

Descriptive statistics were used to summarize baseline characteristics of the study population. Chi-square tests examined variable distributions across groups. Cox proportional hazards models were employed for time-to-event outcomes (CSM and OM). Adjusted probabilities were visualized using margins plots. Statistical significance was determined with a two-tailed alpha level of p < 0.05. All analyses were performed using STATA 16.

## Results

**Table 1** summarizes the baseline demographic and clinical characteristics of 119,937 lung cancer patients, with 47.87% from Texas and 52.12% from California. The population was further divided into pre-ACA (n=60,010; 50.0%) and post-ACA (n=59,927; 49.9%) periods. Pre-ACA, 46.9% of patients were from Texas, and 53.1% were from California; similar proportions were observed post-ACA.

**Table 1.**
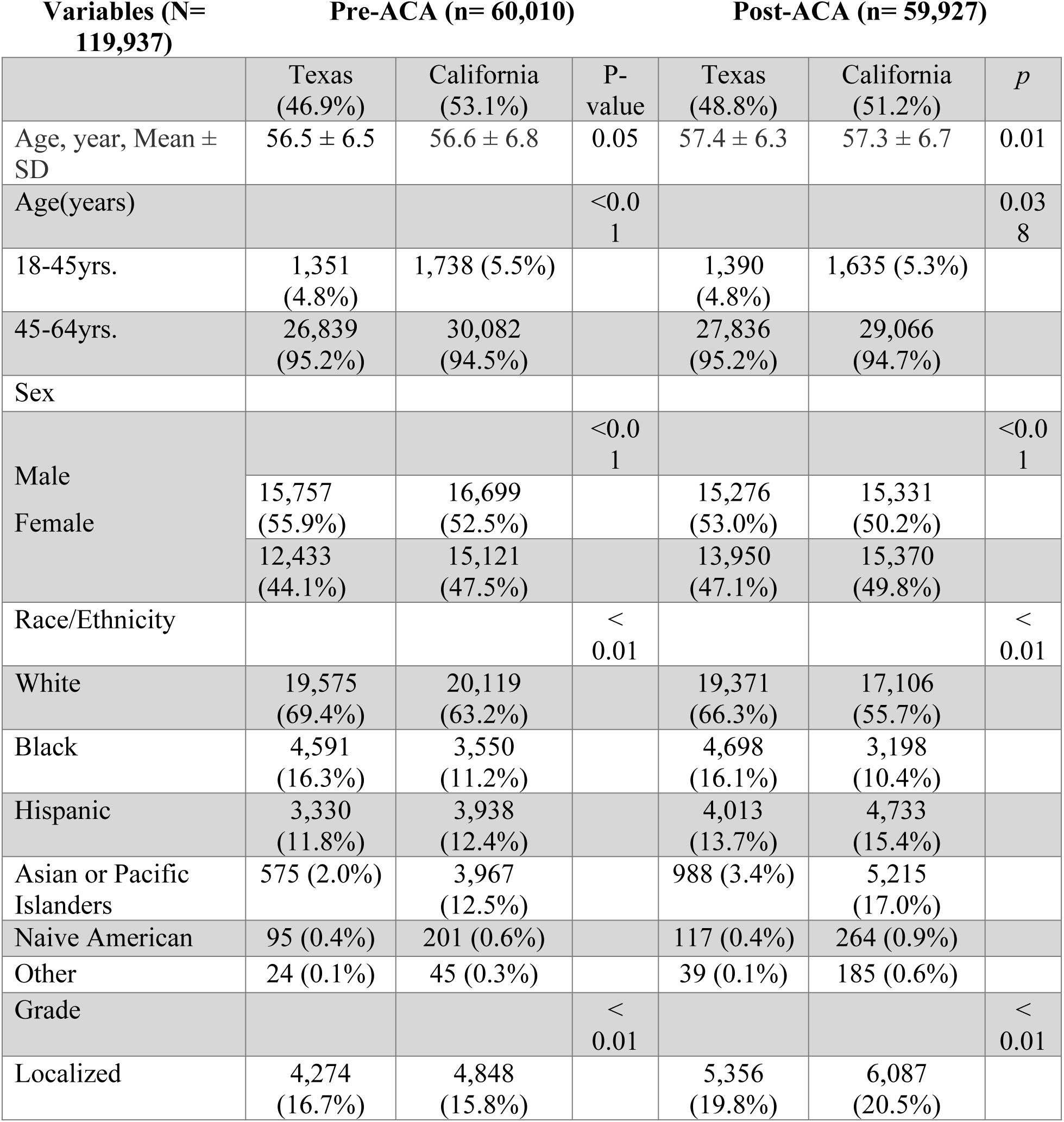
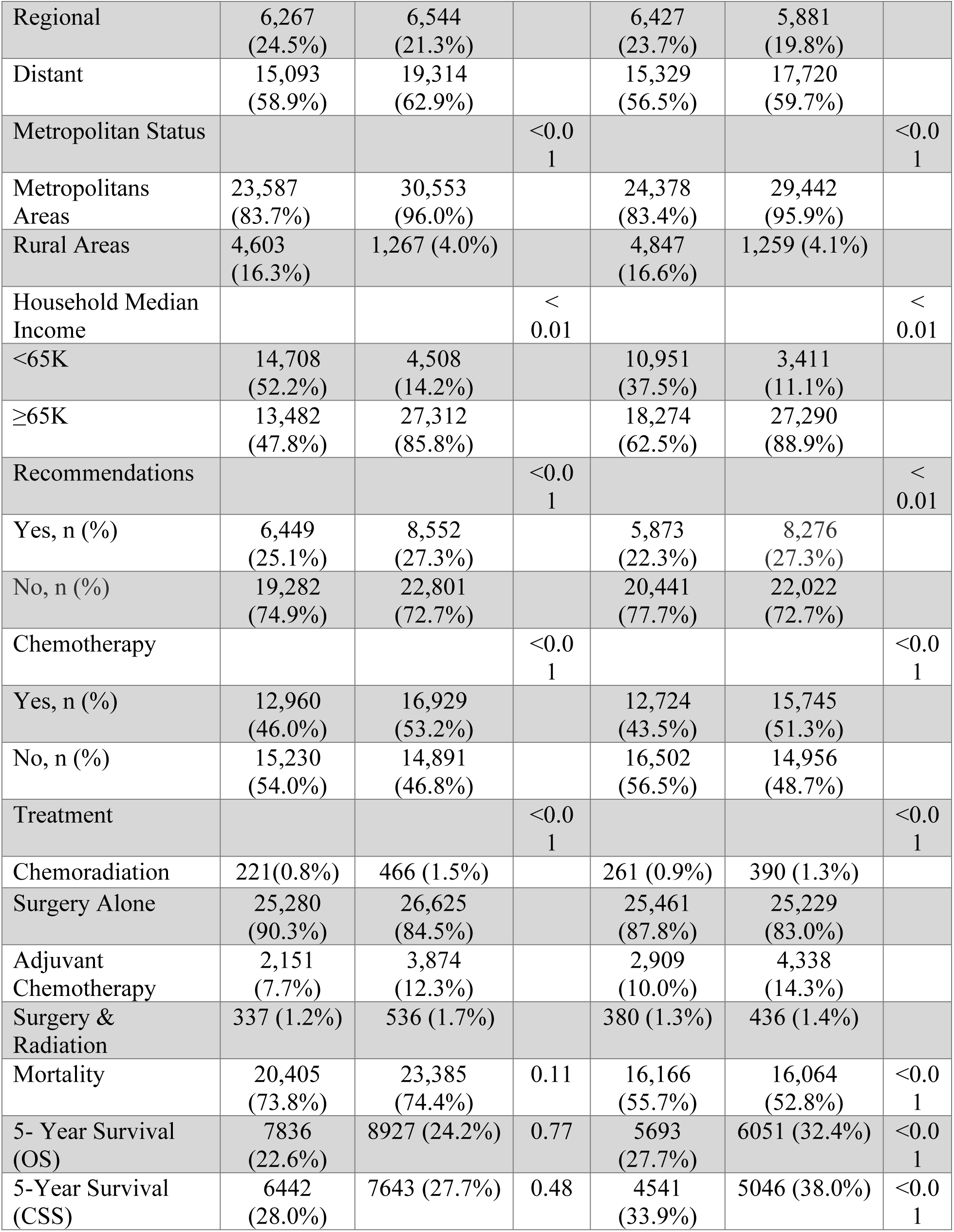
Baseline Demographic and Clinical Characteristics of Patients with Lung Cancer in Texas and California Pre- and Post-ACA Implementation. summarizes the baseline characteristics of cancer patients from California and Texas pre- and post-ACA period. The result shows statistically significant differences in sex, race/ethnicity, household income, and metropolitan status.

Pre-ACA, significant differences were noted between the two states in age, sex, race/ethnicity, household income, and metropolitan status. California had a higher proportion of younger patients aged 18-45 years (5.5% vs. 4.8%, p<0.01), females (47.5% vs. 44.1%, p<0.01), and individuals from racial/ethnic minority groups, including Hispanic (12.4% vs. 11.8%, p<0.01) and Asian/Pacific Islanders (12.5% vs. 2.0%, p<0.01). A greater percentage of patients in California resided in metropolitan areas (96.0% vs. 83.7%, p<0.01) and had a household income ≥$65K (85.8% vs. 47.8%, p<0.01).

Post-ACA, some disparities persisted, but key improvements were noted in both states. The proportion of individuals with distant disease increased in both states but remained marginally higher in California (59.7% vs. 56.5%, p<0.01). California patients had high rates of chemotherapy (51.3% vs. 43.5%, p<0.01) and adjuvant chemotherapy (14.3% vs. 10.0%, p<0.01) compared to their counterparts in Texas. Notably, five-year overall survival (OS) and cancer-specific survival (CSS) improved in both states, with greater gains in California (OS: 32.4% vs. 27.7%, p<0.01; CSS: 38.0% vs. 33.9%, p<0.01). These findings highlight the impact of Medicaid expansion in improving access to care and survival outcomes, particularly for underserved populations in California.

**Table 2** presents the results of the DID analysis comparing CSM among individuals with lung cancer in California (expansion state) versus Texas (non-expansion state, reference) following Medicaid expansion. Overall, CSM significantly decreased in California compared to Texas, with a contrast of −1.12 (95% CI: −1.46 to −0.77, p<0.001). Stratified by income, the reduction was more pronounced among individuals earning less than $65K, with a contrast of −1.26 (95% CI: −1.65 to −0.88, p<0.001), compared to those earning $65K or more, who had a contrast of - 1.11 (95% CI: −1.44 to −0.77, p<0.001). These findings highlight the greater survival benefit of Medicaid expansion for lower-income individuals, emphasizing its role in reducing disparities in lung cancer outcomes.

**Table 2.** Change in CSM among individuals with lung cancer Following Medicaid Expansion: Stratification by Income types. summarizes the results of a Difference-in-Differences analysis comparing lung cancer CSM between Texas (non-expansion state, reference) and California (expansion state) after Medicaid expansion. Significant reductions in CSM were observed across all income groups, with the largest improvement seen among individuals earning <$65K in California compared to their counterparts in Texas.

|  | Contrast | Std. Err. | Z | P>\|z\| | 95%CI |  |
| --- | --- | --- | --- | --- | --- | --- |
| All | -1.12 | 0.176 | -6.34 | <0.001 | -1.46 | -0.77 |
| Income (<\$65K) | -1.26 | 0.196 | -6.43 | <0.001 | -1.65 | -0.88 |
| Income (≥\$65K) | -1.11 | 0.171 | -6.47 | <0.001 | -1.44 | -0.77 |

**Table 3** presents the results of a DID analysis comparing OS among individuals with lung cancer in California versus Texas following Medicaid expansion. Overall, significant reductions in OS were observed in California compared to Texas, with a contrast of −0.81 (95% CI: −1.06 to −0.57, p<0.001). Stratified by income, the reduction in OS was greatest among individuals earning less than $65K, with a contrast of −0.91 (95% CI: −1.18 to −0.64, p<0.001), compared to those earning $65K or more, who had a contrast of −0.78 (95% CI: −1.01 to −0.55, p<0.001). These findings underscore the impact of Medicaid expansion on reducing overall mortality, with a particularly strong benefit for lower-income individuals, further highlighting the role of expanded coverage in addressing healthcare disparities.

**Table 3.** Change in OS among individuals with lung cancer Following Medicaid Expansion: Stratification by Income types. summarizes the results of a Difference-in-Differences analysis comparing lung cancer overall mortality between Texas (non-expansion state, reference) and California (expansion state) after Medicaid expansion. Significant reductions in overall mortality were observed across all income groups, with the largest improvement seen among individuals earning <$65K in California compared to their counterparts in Texas.

|  | Contrast | Std. Err. | Z | P>\|z\| | 95%CI |  |
| --- | --- | --- | --- | --- | --- | --- |
| All | -0.81 | 0.124 | -6.56 | <0.001 | -1.06 | -0.57 |
| Income (<\$65K) | -0.91 | 0.139 | -6.53 | <0.001 | -1.18 | -0.64 |
| Income (≥\$65K) | -0.78 | 0.119 | -6.56 | <0.001 | -1.01 | -0.55 |

**Table 4** summarizes the results of a DID analysis comparing CSM among individuals with lung cancer in California versus Texas after Medicaid expansion, stratified by disease stage at presentation. Overall, CSM significantly decreased in California compared to Texas, with a contrast of −1.12 (95% CI: −1.46 to −0.77, p<0.001). When stratified by disease stage, the magnitude of CSM reduction increased with disease severity.

**Table 4.** Change in CSM among individuals with lung cancer Following Medicaid Expansion: Stratification by Disease Stage. presents the results of a Difference-in-Differences analysis comparing CSM between Texas (non-expansion state, reference) and California (expansion state) after Medicaid expansion. Significant improvements in CSM were observed across all disease stages, with the magnitude of improvement increasing with disease severity. The largest reduction in CSM was observed among individuals with distant metastatic disease, followed by those with regional disease, and the smallest reduction among those with localized disease.

|  | Contrast | Std. Err. | Z | P>\|z\| | 95%CI |  |
| --- | --- | --- | --- | --- | --- | --- |
| All | -1.12 | 0.176 | -6.34 | <0.001 | -1.46 | -0.77 |
| Localized | -0.22 | 0.034 | -6.49 | <0.001 | -0.28 | -0.15 |
| Regional | -0.63 | 0.098 | -6.45 | <0.001 | -0.82 | -0.44 |
| Distant Metastasis | -1.66 | 0.258 | -6.46 | <0.001 | -2.17% | -1.16 |

For patients with localized disease, the contrast was −0.22 (95% CI: −0.28 to −0.15, p<0.001), indicating a smaller but significant reduction in CSM. Among those with regional disease, the reduction was more pronounced, with a contrast of −0.63 (95% CI: −0.82 to −0.44, p<0.001). The largest improvement was observed in individuals presenting with distant metastasis, with a contrast of −1.66 (95% CI: −2.17 to −1.16, p<0.001). These findings highlight the substantial impact of Medicaid expansion on reducing lung cancer mortality, particularly among patients with advanced-stage disease.

**Table 5** presents the results of a DID analysis comparing OS among individuals with lung cancer in California versus Texas following Medicaid expansion, stratified by disease stage at presentation. Overall, OS significantly improved in California compared to Texas, with a contrast of −0.81 (95% CI: −1.06 to −0.57, p<0.001).

**Table 5.** Change in OS among individuals with lung cancer Following Medicaid Expansion: Stratification by Disease Stage. presents the results of a Difference-in-Differences analysis comparing overall mortality between Texas (non-expansion state, reference) and California (expansion state) after Medicaid expansion. Significant improvements in overall mortality were observed across all disease stages, with the magnitude of improvement increasing with disease severity. The largest reduction in overall mortality was observed among individuals with distant metastatic disease, followed by those with regional disease, and the smallest reduction among those with localized disease.

|  | Contrast | Std. Err. | Z | P>\|z\| | 95%CI |  |
| --- | --- | --- | --- | --- | --- | --- |
| All | -0.81 | 0.124 | -6.56 | <0.001 | -1.06 | -0.57 |
| Localized | -0.22 | 0.033 | -6.57 | <0.001 | -0.28 | -0.15 |
| Regional | -0.47 | 0.072 | -6.54 | <0.001 | -0.61 | -0.33 |
| Distant Metastasis | -1.15 | 0.176 | -6.55 | <0.001 | -1.50 | -0.81 |

When stratified by disease stage, the smallest reduction in OS was observed among patients with localized disease, with a contrast of −0.22 (95% CI: −0.28 to −0.15, p<0.001). Patients with regional disease experienced a greater reduction, with a contrast of −0.47 (95% CI: −0.61 to −0.33, p<0.001). The largest improvement was seen in individuals presenting with distant metastasis, with a contrast of −1.15 (95% CI: −1.50 to −0.81, p<0.001).

**Table 6** presents the results of a DID analysis comparing CSM among Medicaid-eligible individuals with lung cancer in California versus Texas following Medicaid expansion, stratified by race/ethnicity. Overall, CSM significantly decreased in California compared to Texas, with a contrast of −1.12 (95% CI: −1.46 to −0.77, p<0.001).

**Table 6.** Change in CSM among individuals with lung cancer following Medicaid Expansion: Stratification by Race/Ethnicity. presents the results of a Difference-in-Differences analysis comparing CSM between Texas (non-expansion state, reference) and California (expansion state) after Medicaid expansion. Significant reductions in CSM were observed across all racial/ethnic groups, with the largest reduction seen among Black individuals, followed by White and Hispanic individuals.

|  | Contrast | Std. Err. | Z | P>\|z\| | 95%CI |  |
| --- | --- | --- | --- | --- | --- | --- |
| All | -1.12 | 0.176 | -6.34 | <0.001 | -1.46 | -0.77 |
| White | -1.16 | 0.182 | -6.34 | <0.001 | -1.51 | -0.80 |
| Black | -1.25 | 0.198 | -6.32 | <0.001 | -1.64 | -0.86 |
| Hispanic | -1.02 | 0.162 | -6.32 | <0.001 | -1.34 | -0.71 |
| Asians/Pacific Islander | -0.82 | 0.130 | -6.33 | <0.001 | -1.08 | -0.57 |

Stratified by race/ethnicity, the largest reduction in CSM was observed among Black individuals, with a contrast of −1.25 (95% CI: −1.64 to −0.86, p<0.001), followed by White individuals (−1.16; 95% CI: −1.51 to −0.80, p<0.001) and Hispanic individuals (−1.02; 95% CI: −1.34 to −0.71, p<0.001). Asian/Pacific Islanders experienced the smallest reduction in CSM, with a contrast of **-** 0.82 (95% CI: −1.08 to −0.57, p<0.001). These findings highlight the significant impact of Medicaid expansion in reducing lung cancer mortality across all racial/ethnic groups, with the most pronounced benefits observed among Black individuals.

**Table 7** presents the results of the DID analysis comparing overall mortality among individuals with lung cancer in California vs. Texas following Medicaid expansion, stratified by race/ethnicity. The Overall mortality significantly decreased in California compared to Texas, with a difference of −0.81 (95% CI: −1.06 to −0.57, p<0.001).

**Table 7.** Change in OS among individuals with lung cancer Following Medicaid Expansion: Stratification by Disease Stage. presents the results of a Difference-in-Differences analysis comparing overall mortality between Texas (non-expansion state, reference) and California (expansion state) after Medicaid expansion. Significant reductions in overall mortality were observed across all racial/ethnic groups, with the largest reduction seen among Black individuals, followed by White and Hispanic individuals.

|  | Contrast | Std. Err. | Z | P>\|z\| | 95%CI |  |
| --- | --- | --- | --- | --- | --- | --- |
| All | -0.81 | 0.124 | -6.56 | <0.001 | -1.06 | -0.57 |
| White | -0.84 | 0.128 | -6.56 | <0.001 | -1.09 | -0.59 |
| Black | -0.91 | 0.140 | -6.54 | <0.001 | -1.19 | -0.64 |
| Hispanic | -0.76 | 0.117 | -6.54 | <0.001 | -0.99 | -0.53 |
| Asian or Pacific is | -0.59 | 0.090 | -6.54 | <0.001 | -0.76 | -0.41 |

Stratified by race/ethnicity, the largest reduction in overall mortality was observed among Black individuals, with a contrast of −0.91 (95% CI: −1.19 to −0.64, p<0.001), followed by White individuals (−0.84; 95% CI: −1.09 to −0.59, p<0.001) and Hispanic individuals (−0.76; 95% CI: - 0.99 to −0.53, p<0.001). Asian/Pacific Islanders experienced the smallest reduction in overall mortality, with a contrast of −0.59 (95% CI: −0.76 to −0.41, p<0.001).

## Discussion

We observed a substantial reduction in both cancer-specific mortality and overall deaths in California compared to Texas following the implementation of Medicaid expansion under the Affordable Care Act (ACA). Individuals in California experienced significant improvements in survival, encompassing both cancer-specific and overall survival, when compared to those in Texas, a non-expansion state. Notably, this survival benefit was consistent across income groups, racial/ethnic categories, and disease stages at presentation. These findings underscore the broad and equitable impact of Medicaid expansion on improving cancer outcomes.

### Income-Based Survival Disparities

Individuals earning less than $65,000 annually demonstrated the most pronounced improvement in survival after Medicaid expansion. However, individuals earning more than $65,000 also experienced significant survival gains compared to their counterparts in non-expansion Texas. This disparity may be attributable to improved access to health insurance among lower-income groups in California, facilitating early diagnosis, timely treatment, and access to advanced therapies[16, 25, 26]. Medicaid expansion has likely reduced financial barriers to care, particularly for high-cost interventions such as chemotherapy, targeted therapies, and supportive care for low-income populations[27, 28].

### Stage-Based Survival Trends

Interestingly, the largest survival improvement was observed among individuals with advanced-stage disease (distant metastasis), followed by regional disease, with the least improvement seen in localized disease. This trend can be attributed to several interrelated factors. Pre-expansion, individuals with distant metastasis in non-expansion states like Texas faced significant barriers to accessing high-cost, complex care, which contributed to poorer outcomes[29]. The implementation of Medicaid expansion in California addressed these disparities by improving access to advanced therapies, resulting in marked reductions in mortality among this population[16, 17, 30].

Comprehensive coverage for advanced disease also played a crucial role in these survival improvements. Treatments for advanced-stage cancer, such as targeted therapy and immunotherapy, are often prohibitively expensive for uninsured or underinsured patients. Medicaid expansion in California ensured that such high-cost interventions became accessible to a broader group of patients, significantly improving outcomes[31]. Additionally, the expansion increased access to palliative care services, which are known to enhance both quality of life and survival for patients with advanced cancer, further contributing to the observed benefits[31, 32].

In contrast, patients with localized disease demonstrated smaller survival improvements, largely due to a “ceiling effect.” These patients typically already have excellent prognoses due to early detection and the availability of effective, often curative treatments. As a result, there was limited room for further mortality reduction with Medicaid expansion. Moreover, insurance status may have a relatively minor impact on outcomes for early-stage disease compared to advanced stages, where continuous and specialized care is more critical[33, 34].

Another factor influencing these trends is selection bias and stage migration. In non-expansion states, delays in care due to lack of insurance often result in diagnoses at more advanced stages[21, 35–37]. Conversely, Medicaid expansion in California likely facilitated earlier diagnoses and improved staging accuracy, contributing to better outcomes. This phenomenon may have also resulted in a more accurate categorization of advanced-stage patients in expansion states, making them more comparable to regional-stage patients in non-expansion states and thereby reducing observed mortality differences[30]. Together, these factors illustrate the multifaceted impact of Medicaid expansion on cancer survival outcomes across different stages of disease.

### Race/Ethnicity-Based Survival Disparities

The survival improvement was observed across all major racial/ethnic groups, including Black, White, Hispanic, and Asian/Pacific Islander individuals. Although our study lacked sufficient sample sizes to draw robust conclusions for Native American and other racial groups, it is noteworthy that Black individuals exhibited the highest survival benefit, followed closely by Whites and Hispanics. These findings align with the hypothesis that Medicaid expansion disproportionately benefits historically marginalized populations by addressing long-standing disparities in access to care[38, 39]. The survival gains among racial minorities may also reflect improved access to culturally competent care and enhanced utilization of preventive and therapeutic services[38, 39].

### Healthcare System and Policy Implications

The improved outcomes observed in California are attributable not solely to the Medicaid expansion but also to systemic factors that enhance the overall quality of healthcare delivery. California’s robust healthcare infrastructure, which includes well-established provider networks and advanced cancer care facilities, has likely played a pivotal role in amplifying the benefits of Medicaid expansion. Additionally, the state’s implementation of comprehensive cancer programs has contributed to improved outcomes, particularly for patients requiring complex and resource-intensive treatments.

Expansion states, including California, may also benefit from the adoption of complementary public health initiatives aimed at addressing broader determinants of health. These initiatives could include cancer screening programs, patient navigation services, and efforts to reduce disparities in access to care. By targeting high-risk populations and enhancing early detection, these programs likely synergized with Medicaid expansion to deliver substantial survival benefits. Collectively, these systemic and policy-driven factors highlight the critical need for holistic approaches that integrate policy reforms with investments in healthcare infrastructure and public health programs to achieve equitable and sustainable improvements in cancer outcomes.

### Limitations

Our study has several limitations that should be acknowledged. First, we employed a complete case analysis, excluding patients with missing data. This approach may introduce selection bias and limit the generalizability of our findings, as exclusions due to missing data may not be random and could potentially affect the validity of our results. Second, the absence of critical variables, such as detailed comorbidity information, baseline insurance status, and patient-reported outcomes, limits our ability to control for pre-existing health conditions and other unmeasured confounders that might influence survival outcomes. These missing data elements are particularly pertinent when analyzing the complexities of cancer treatment and progression.

Third, while the SEER registry provides a robust dataset for cancer research, it lacks information on systemic treatment details, such as access to targeted therapies or immunotherapies, which are critical for lung cancer management. Additionally, SEER does not capture certain socioeconomic and behavioral factors, such as smoking history, which are particularly relevant to lung cancer and could serve as significant confounders. Fourth, although lung cancer is not an indolent disease like prostate cancer, and the survival times are relatively shorter, our study may still be limited by the follow-up period, which restricts our ability to fully assess the long-term impact of Medicaid expansion. Future research with extended follow-up periods and the inclusion of additional covariates may provide a more comprehensive understanding of these effects.

Finally, while our analysis adjusted for race, income, and metropolitan status, residual confounding remains a concern. Factors such as variations in healthcare utilization, differences in provider practices, and patient quality of life, which are not captured in the SEER registry, may influence the observed outcomes despite statistical adjustments. Addressing these limitations in future studies by incorporating datasets with richer clinical and socioeconomic variables could enhance the robustness and applicability of the findings.

## Conclusion

This study underscores the transformative effect of Medicaid expansion in significantly reducing cancer-specific and overall mortality, with the greatest benefits observed among vulnerable populations. The pronounced improvements in survival for individuals with advanced-stage disease, lower-income groups, and racial minorities highlight the vital importance of equitable healthcare policies in addressing longstanding disparities in access to care and outcomes. These findings demonstrate the potential of Medicaid expansion to bridge critical gaps in healthcare delivery and improve population health. Future research should focus on evaluating the long-term effects of Medicaid expansion and examining the interplay between policy changes and systemic factors to further enhance its impact and sustainability.

## Data Availability

All relevant data are within the manuscript and its Supporting Information files.

## Acknowledgments

The authors would like to acknowledge the Clive O. Callender Outcomes Research Center at the Howard University College of Medicine for its support in facilitating this research.

